# Higher case fatality rate among obstetric patients with COVID-19 in the second year of pandemic in Brazil: do new genetic variants play a role?

**DOI:** 10.1101/2021.05.06.21256651

**Authors:** Maíra L S Takemoto, Marcos Nakamura-Pereira, Mariane O Menezes, Leila Katz, Roxana Knobel, Melania M R Amorim, Carla B Andreucci

**Affiliations:** São Paulo State University (UNESP), Medical School of Botucatu. Address: Av. Prof. Montenegro, s/n - Botucatu - SP, Brazil. Postal code: 18618-687; National Institute for Women, Children and Adolescents Health Fernandes Figueira, Oswaldo Cruz Foundation. Address: Av. Rui Barbosa, 716 – Flamengo. Rio de Janeiro - RJ, Brazil. Postal code: 22250-020; Professor Fernando Figueira Institute of Integral Medicine (IMIP). Address: R. dos Coelhos, 300 - Boa Vista, Recife - PE, Brazil. Postal code: 50070-902; Federal University of Santa Catarina (UFSC), Department of Gynecology and Obstetrics. Address: R. Eng. Agronômico Andrei Cristian Ferreira, s/n - Trindade, Florianópolis - SC, Brazil. Postal code: 88040-900; Federal University of Sao Carlos (UFSCAR), Department of Medicine. Address: Rodovia Washington Luis, km 235 - São Carlos - SP, Brazil. Postal code: 13565-905

## Abstract

**Background:** In Brazil, a 20% increase in maternal mortality rate due to COVID-19 is projected for 2020. On January 4, 2021, the P.1 SARS-CoV-2 genetic variant was firstly identified in the country and recent data has indicated an association with higher hospitalization rates and mortality. The impact of P.1 variant in the obstetric population remains unclear.

**Methods:** We carried out a preliminary analysis of sociodemographic and clinical characteristics of COVID-19 confirmed maternal deaths (between 10-50 years old) comparing cases reported to the Brazilian official severe acute respiratory syndrome (SARS) surveillance system (SS) in 2020 with those from 2021 (until April 12, 2021). This preliminary analysis employed methods described in previous reports from our group.

**Results:** 803 maternal deaths out of 8,248 COVID-19 maternal SARS cases with a recorded outcome were reported to the SARS-SS since March 2020. Case fatality rate was significantly higher in 2021 (15.6% vs 7.4%). The first three months of 2021 already account for 46.2% of all deaths occurred in the 13-months analysed period. COVID-19 fatal cases from 2021 had a lower proportion of at least one risk factor or comorbidity as compared to 2020 but had a higher frequency of obesity. There were no significant differences in terms of age, type of residence area (urban, rural, or peri-urban), type of funding of the notification unit (public vs. private), COVID-19 diagnostic criteria, pregnancy status (pregnancy or postpartum), cardiovascular disease or diabetes. The proportion of hospitalization, ICU admission, and respiratory support before death was also not significantly different.

**Conclusion:** Case fatality rate was increased in the three first months of 2021 when compared to 2020. Once variables related to health care access and demographics are not significantly different and women seem to be healthier in the 2021 sample, such difference may be related to the circulation of more aggressive genetic variants in the country.

---

The COVID-19 pandemic inflicted an overwhelming burden upon the Brazilian obstetric population^1,2^. It is estimated that it has already been responsible for a 20% increase in maternal mortality rate in the country in 2020^3^. Analysis from the Brazilian official severe acute respiratory syndrome (SARS) surveillance system (SS) observed worse outcomes in obstetric patients with older age and comorbidities, in concordance with findings for the general population^4^. Socioeconomic factors that reflect barriers to access healthcare seems to have adversely impacted COVID-19-related maternal mortality in Brazil^5^. Considering maternal death as a sentinel event to assess health in a population, the Brazilian historically overloaded and underfunded healthcare system collapse is likely playing a role in such tragedy.

On January 4, 2021, the P.1 SARS-CoV-2 genetic variant was firstly identified in Brazil. Since the description of the strain, epidemiology of COVID-19 infection in the general Brazilian population seems to have changed. Published reports have shown greater community transmissibility of the P.1 variant, accompanied by increase in mortality rates, particularly in younger people^6,7^. In February 2021, a study conducted in an area with exponentially increasing hospitalisation rate in the South region identified the P.1 variant in 24 out of 27 SARS-CoV-2 samples.^6^

To examine if the profile of COVID-19 maternal deaths is also modified after the new variant identification, we carried out a preliminary analysis of sociodemographic and clinical characteristics of COVID-19 confirmed maternal deaths comparing cases reported to the SARS-SS in 2020 and 2021 (in both public and private hospitals, until April 12, 2021). This preliminary analysis follows methods described in previous reports from our group^4,5^.

Findings by year of notification in the SARS-SS are displayed in Table 1. Case fatality rate was significantly higher in 2021 (15.6% vs 7.4%) with 803 maternal deaths out of 8,248 COVID-19 SARS maternal cases with a recorded outcome since March 2020. The first three months of 2021 already account for 46.2% of all deaths occurred in the 13-months analysed period. At the same time, the number of SARS notifications in 2021 regardless of the outcome is only 34.4% of the total number for the entire period (3,474 out of 10,080, data not shown).

**Table 1.** Comparison of fatal cases of COVID-19 during pregnancy or postpartum in Brazil by year of notification to SARS-SS (2020 vs. 2021)

|  | All cases |  | 2020 |  | 2021 |  | p-value* |
| --- | --- | --- | --- | --- | --- | --- | --- |
|  | N | % | n | % | n | % |  |
| Case fatality rate | 803 | 9.7 | 432 | 7.4 | 371 | 15.6 | <0.001 |
| Age group |  |  |  |  |  |  |  |
| 10-19 | 28 | 3.5 | 21 | 4.9 | 7 | 1.9 | 0.121 |
| 20-34 | 487 | 60.6 | 258 | 59.7 | 229 | 61.7 |  |
| 35-44 | 268 | 33.4 | 144 | 33.3 | 124 | 33.4 |  |
| 45+ | 20 | 2.5 | 9 | 2.1 | 11 | 3.0 |  |
| Ethnicity/Skin color |  |  |  |  |  |  |  |
| White | 234 | 29.8 | 108 | 26.2 | 126 | 34.0 | 0.027 |
| Black | 55 | 7.0 | 39 | 9.4 | 16 | 4.3 |  |
| Yellow | 7 | 0.9 | 5 | 1.2 | 2 | 0.5 |  |
| Brown | 412 | 52.6 | 222 | 53.8 | 190 | 51.2 |  |
| Indigenous | 6 | 0.8 | 3 | 0.7 | 3 | 0.8 |  |
| Missing | 70 | 8.9 | 36 | 8.7 | 34 | 9.2 |  |
| Geographic region |  |  |  |  |  |  |  |
| North | 139 | 17.3 | 61 | 14.1 | 78 | 21.0 | <0.001 |
| Northeast | 212 | 26.4 | 137 | 31.7 | 75 | 20.2 |  |
| Mid-West | 75 | 9.3 | 37 | 8.6 | 38 | 10.3 |  |
| Southeast | 300 | 37.4 | 172 | 39.8 | 128 | 34.5 |  |
| South | 77 | 9.6 | 25 | 5.8 | 52 | 14.0 |  |
| Type of residence area |  |  |  |  |  |  |  |
| Urban | 652 | 81.2 | 352 | 81.5 | 300 | 80.9 | 0.207 |
| Rural | 64 | 8.0 | 31 | 7.2 | 33 | 8.9 |  |
| Peri-urban | 4 | 0.5 | 4 | 0.9 | 0 | 0.0 |  |
| Missing | 83 | 10.3 | 45 | 10.4 | 38 | 10.2 |  |
| Funding of SARS notification unit |  |  |  |  |  |  |  |
| Public | 658 | 81.9 | 352 | 81.5 | 306 | 82.5 | 0.714 |
| Private | 145 | 18.1 | 80 | 18.5 | 65 | 17.5 |  |
| Diagnostic criteria |  |  |  |  |  |  |  |
| Laboratory | 729 | 90.8 | 393 | 91.0 | 336 | 90.6 | 0.142 |
| Clinical-Epidemiological | 13 | 1.6 | 4 | 0.9 | 9 | 2.4 |  |
| Clinical | 15 | 1.9 | 11 | 2.5 | 4 | 1.1 |  |
| Clinical-Imaging | 33 | 4.1 | 16 | 3.7 | 17 | 4.6 |  |
| Missing | 13 | 1.6 | 8 | 1.9 | 5 | 1.3 |  |
| Pregnancy status |  |  |  |  |  |  |  |
| Postpartum | 301 | 37.5 | 169 | 39.1 | 132 | 35.6 | 0.301 |
| Pregnant | 502 | 62.5 | 263 | 60.9 | 239 | 64.4 |  |
| **No risk factor or comorbidity** | 439 | 54.7 | 220 | 50.9 | 219 | 59.0 | **0.021** |
| **Cardiovascular disease** | 109 | 13.6 | 63 | 14.6 | 46 | 12.4 | 0.510 |
| **Diabetes** | 103 | 12.8 | 63 | 14.6 | 40 | 10.8 | 0.363 |
| **Obesity** | 109 | 13.6 | 51 | 11.8 | 58 | 15.6 | **0.041** |
| **Inpatient admission** | 777 | 96.8 | 421 | 97.5 | 356 | 96.0 | 0.106 |
| **ICU admission** | 555 | 69.1 | 296 | 68.5 | 259 | 69.8 | 0.769 |
| **Respiratory support** |  |  |  |  |  |  |  |
| Yes, invasive | 481 | 59.9 | 250 | 57.9 | 231 | 62.3 | 0.195 |
| Yes, non-invasive | 161 | 20.0 | 81 | 18.8 | 80 | 21.6 |  |
| No | 161 | 20.0 | 101 | 23.4 | 60 | 16.2 |  |
| **Time from symptom onset to inpatient admission – median (95% CI) (n=771)** | 5 | (5-6) | 5 | (4-5) | 6 | (5-6) | **<0.001** |
| **Time from hospital admission to ICU admission – median (95% CI) (n=543)** | 0 | (0-0) | 0 | (0-0) | 0 | (0-0.5) | 0.141 |
| **Length of ICU stay – median (95% CI) (n=338)** | 10 | (9-11) | 11 | (9.3-13.7) | 9 | (7-10.7) | 0.269 |
| **Time from hospital admission to death – median (95% CI) (n=771)** | 11 | (10-12) | 11 | (10-12) | 11 | (9.1-12) | 0.659 |

CI, confidence interval; ICU, intensive care unit; SARS, severe acute respiratory syndrome; \*Chi-square p-values except for the continuous variables (Two-sample Mann-Whitney test)

There was no significant difference in terms of age, type of residence area, type of funding of the notification unit, COVID-19 diagnostic criteria, distribution of pregnancy status, presence of cardiovascular disease or diabetes. The proportion of hospitalization, ICU admission, and respiratory support before death was also not significantly different.

However, the proportion of white women among cases from 2021 was slightly higher, as well as missing data for ethnicity was reduced. There was a reduction in the Northeast region participation in the overall number of deaths in 2021, with a simultaneous increase in South region participation. Fatal cases for 2021 were also less likely to have at least one risk factor (59% had no risk factor or comorbidity) and more likely to be obese. Median time from symptoms onset to inpatient admission due to SARS was slightly longer in 2021, but time from hospital admission to ICU admission, from admission to death, and length of ICU stay before death did not differ. Noteworthy, median time from hospital admission to ICU time was zero days in both years, implying that patients have been admitted already under severe clinical conditions regardless of year of occurrence.

We believe that health system collapse cannot be ruled out as one of the possible reasons for the increase in the case-fatality rate. We examined hospitalized cases with SARS only and barriers to access proper treatment in earlier stages of the disease may have worsen in 2021. For instance, the beginning of 2021 in Brazil was marked by news of collapsed private hospitals all over the country, which did not happen in 2020^8^. However, our preliminary data seems to indicate that in 2021, healthier Brazilian women are dying despite the relative stability of other risk factors. The increased participation of white women as well as from wealthier geographic regions (markedly the South region) reinforces this hypothesis. Our findings are in line with recent reports from the UK indicating that pregnant women may be more affected in the second pandemic wave^9,10^.

Considering these findings, it is reasonable to believe that new factors have been introduced in the scenario and the impact of the new coronavirus variants may be one of them. The topic should be further studied warranting the inclusion of the obstetric population in genomic surveillance of SARS-CoV-2 in Brazil. Once effective pandemic containment measures including vaccination and mass testing are still not consistently adopted in the country, we are unable to foresee an end for this calamity.

We declare no competing interests.

## Data Availability

The data that support the findings of this study were openly available before the initiation of the study and can be accessed without application

https://opendatasus.saude.gov.br/dataset/bd-srag-2021/resource/42bd5e0e-d61a-4359-942e-ebc83391a137

